# Mapping the uneven landscape: A geospatial analysis of breast cancer incidence and mammography access across Illinois (2017-2021)

**DOI:** 10.1101/2025.06.03.25328939

**Authors:** Waheed O. Ogunwale, Oluwatomi A. Ajao, Olufisayo E. Ogunbayo

## Abstract

Breast cancer continues to place a significant burden on women’s health in the United States, but that burden is not shared equally across regions. This study looks at how breast cancer incidence and access to mammography services differ across Illinois, using data from 2017 to 2021. By analyzing county-level figures on breast cancer cases, facility availability, and social indicators, we identified patterns using spatial tools like ArcGIS Pro, GeoDa, and statistical modeling in R.

Our analysis showed that the risk of being diagnosed with breast cancer varies considerably depending on where a woman lives. Some counties reported incidence rates more than three times higher than others. Meanwhile, mammography facility density varied just as widely, suggesting serious gaps in healthcare infrastructure. Spatial analysis confirmed that high-incidence areas often cluster together, particularly in central Illinois. However, these high-burden counties did not always lack screening centers, pointing to the influence of other factors beyond access alone.

Multivariable models showed that counties with higher education levels tended to report more diagnoses, likely due to better screening participation. Conversely, more facilities per capita were associated with slightly lower incidence, possibly reflecting earlier detection and prevention. Race and poverty did not show significant associations once spatial and statistical adjustments were made.

These findings suggest the need for more than just increasing the number of screening centers. Effective solutions will require investing in education, improving outreach, and addressing broader systemic barriers to care. Public health strategies can become more targeted, equitable, and effective by considering geography and local context.

## Background

Breast cancer continues to rank as the most commonly diagnosed cancer among women in the United States, and it continues to rank among the leading causes of cancer-related death despite substantial advances in screening and treatment [1,2]. Although national mortality has declined, those aggregate gains mask pronounced geographic and sociodemographic disparities. Rural counties, for example, report higher proportions of late-stage presentation and poorer survival independent of tumor biology or treatment advances [3,4]. Several extensive surveillance studies attribute these gaps, in part, to uneven access to screening, delayed diagnostic work-up, and structural barriers embedded in local health systems [5,6].

Spatial access to mammography is a pivotal determinant of early detection. Women who must travel farther for imaging are less likely to obtain guideline-concordant screening, particularly if they live in low-income areas or lack insurance coverage [7]. Prior work has documented a substantial travel burden to breast imaging in the Mountain West, Appalachia, and the Deep South [8,9]. However, Midwestern states remain understudied even though they blend dense metropolitan hubs and expansive rural regions. Illinois provides a compelling case in point.

Chicago’s Cook County hosts a high concentration of radiology services, whereas large stretches of the state remain agricultural and sparsely populated. Despite this dichotomy, the spatial distribution of mammography resources and county-level breast-cancer incidence has not been comprehensively mapped since digital breast tomosynthesis became standard.

Early evidence suggests that facility supply alone does not fully explain observed disparities. A 2023 Delaware-based catchment area study showed that expanding mammography facility availability in targeted regions helped shorten diagnostic delays among underserved populations, independent of socioeconomic influences [10]. Conversely, a detailed analysis of breast cancer screening disparities showed that a dynamic interplay of multiple factors continues to perpetuate inequitable access despite several policies and interventions, indicating that additional context such as socioeconomic status, environmental exposures, trust in the health system, or scheduling logistics may modulate the impact of raw facility numbers [7]. Population-based breast cancer counts from the Illinois State Cancer Registry [11] coupled with the audited registry of certified mammography units from the US Food and Drug Administration (FDA) [12], Illinois geospatial data from the Illinois Department of Transportation [13], granular sociodemographic covariates from the American Community Survey (ACS) [14] enables a more rigorous assessment of spatial relationships.

Two additional methodological gaps deserve attention. A persistent statistical challenge is the tight linkage among race, income, and educational attainment. When these factors move in tandem, regression models can carry inflated error terms, masking the distinct influence of each indicator. Williams et al. observe that race and socioeconomic factors frequently capture overlapping dimensions of structural disadvantage [15]. Principal component analysis (PCA) offers one solution by distilling correlated measures into orthogonal indices, yet few breast-cancer mapping studies have taken this approach. Spatial autocorrelation violates the independence assumption of ordinary least squares (OLS) and can bias coefficient estimates. Spatial-lag or conditional autoregressive models capture the contagious nature of disease risk across county borders, but they have seldom been paired with over-dispersed count models such as negative binomial regression [16].

Illinois is also policy-relevant. The state expanded Medicaid under the Affordable Care Act and invested in mobile mammography units to reach underserved regions [17,18]. Administrative data reveal increased screening among previously uninsured women, yet it remains unclear whether new capacity has translated into a lower county-level burden or shifted utilization toward urban centers [19]. Identifying counties with high incidence despite ample facilities could flag deeper obstacles like transport, health literacy, or environmental hazards. However, spotting low-incidence but scanner-scarce areas would justify the placement of additional units.

The present study addresses these gaps by integrating verified facility data, population-based cancer counts, and detailed sociodemographic indicators into a spatial analytical framework. We first map county-level breast-cancer incidence from 2017 to 2021 and mammography capacity in 2021, using FDA records for facilities and IDPH registry counts for cases. We then quantify global and local spatial clustering to determine whether high-incidence counties form coherent corridors. Next, we model the association between facility density and incidence rate through ordinary least squares, apply spatial-lag correction, and transition to a principal-component-adjusted negative binomial model to accommodate over-dispersion. This multistep approach allows us to isolate the role of screening infrastructure while accounting for interconnected social determinants. Finally, we examine residual hot spots to highlight areas where additional factors, environmental exposures, delayed follow-up, or cultural barriers, may still operate.

By analyzing Illinois, a state with robust cancer surveillance and heterogeneous geography, the present work advances the understanding of how spatial access and sociodemographic composition interact to shape breast-cancer burden. The findings aim to guide health departments, hospital networks, and community organizations in allocating resources more equitably. Mapping the uneven landscape of breast-cancer incidence and mammography access thus provides an empirically grounded template for narrowing persistent gaps in early detection and outcomes.

## Methods

This cross-sectional ecological study utilized publicly available secondary data to investigate breast cancer incidence and mammography facility access across all 102 counties in Illinois. The analytical period covered 2017 to 2021, aligning with the most recent five-year release from the Illinois State Cancer Registry [11]. Ethical approval was not required since all data were de-identified and aggregated by nature.

Breast cancer case counts included in situ (non-invasive or early stage) and invasive diagnoses among women and were retrieved from the Illinois Department of Public Health (IDPH) [11]. To stabilize year-to-year variability, total case counts per county over five years were standardized by the average female population aged 40 to 74 for the same period, consistent with surveillance best practices [1,2]. This age band aligns with breast cancer screening guidelines from the US Preventive Services Task Force (USPSTF) and the Centers for Disease Control and Prevention (CDC) [6,20].

Population data were sourced from the US Census Bureau’s 2017-2021 American Community Survey 5-year estimates [14]. Average population figures were computed by summing annual estimates from 2017 to 2021 and dividing by five. Sensitivity testing showed minimal deviation when the 2019 midpoint population was used instead, with over 99% of counties showing less than a 5% difference in incidence rates.

Mammography facility data were obtained from the FDA’s Mammography Quality Standards Act database as of November 2021 [12]. Each facility’s address was batch-geocoded using the US Census Geocoder and manually verified for unmatched cases. Spatial joins with the 2024 Illinois county shapefile from the Department of Transportation assigned facilities to counties [13]. Facility density was computed as the number of certified centers per 100,000 women aged 40–74. Facility locations were also visualized to account for geographic disparities using kernel density estimations in ArcGIS Pro 3.4 [21].

Sociodemographic indicators, including poverty rate, educational attainment (percentage of the population with a bachelor’s degree or higher), and racial composition (percentage of the African American, Latino, and non-Hispanic White), were extracted from the ACS [14]. These variables were normalized to proportions to facilitate model interpretation. Race and poverty have historically been linked to disparities in cancer outcomes [22], yet their independent effects remain difficult to isolate due to collinearity [23].

Spatial incidence and facility density patterns were assessed using global and local indicators. Global Moran’s I evaluated spatial autocorrelation at the state level using a 200 km fixed-band inverse distance spatial weights matrix [24,25]. Local Indicators of Spatial Association (LISA) were applied to detect county-level hotspots and cold spots of incidence and service access [26]. Maps were rendered using natural breaks classification in ArcGIS Pro, which optimizes inter-class variance [21].

OLS regression was used to model county-level breast cancer incidence as a function of facility density and sociodemographic covariates. Multicollinearity was assessed using variance inflation factors (VIF), with a threshold of >10 indicating problematic redundancy [27]. Given the high correlations among race variables (Spearman ρ > 0.80), PCA was performed to derive a composite race index consistent with established dimensionality reduction strategies in public health research [28].

Residual diagnostics from the OLS model revealed significant spatial autocorrelation (Moran’s I p < 0.001), suggesting spatial dependence. To address this, a spatial lag model was estimated using GeoDa 1.20, incorporating a spatially lagged dependent variable to account for spillover effects between neighboring counties [29]. The weights matrix for this model was constructed using first-order queen contiguity [30].

Additionally, count-based models were implemented, given the nature of cancer cases. A Poisson model was first estimated but rejected due to substantial overdispersion (dispersion ratio > 30). A negative binomial regression model was fitted and adjusted for PCA-transformed covariates using the MASS package in R. This model demonstrated a better fit, as evidenced by lower AIC and deviance statistics [31,32].

Sensitivity analysis was conducted using an alternate population denominator [33,34] to ensure robustness. Results were compared across population denominators (five-year average population to the 2019 midpoint), and coefficients remained stable (<5% fluctuation), supporting the internal validity of findings.

All data cleaning, statistical modeling, and visualization were performed using RStudio 2024.12.1 and ArcGIS Pro 3.4. The tidyverse suite facilitated data management [35], while tmap and ggplot2 were used for graphics aligned with World Health Organization (WHO) data communication standards [36]. The full codebase and annotated data dictionary are publicly available via Zenodo [37] to promote transparency and reproducibility.

## Results

Breast cancer incidence in Illinois is geographically uneven. Over the 2017–2021 period, the statewide mean was 2,274 cases per 100,000 women aged 40–74 years, yet county rates spanned a four-fold range, from 1,074 in the least-affected jurisdiction to 3,552 in the most affected (Fig. 1; Table 1). The middle half of counties alone showed a swing of nearly nine hundred cases, indicating that large swings are not limited to a handful of statistical outliers. Screening infrastructure displayed greater dispersion. While Illinois averaged approximately nineteen certified mammography units per 100,000 women, county values ranged from a mobile scanner to more than seventy fixed sites, producing a coefficient of variation of 94 percent (Table 1).

**Fig 1.**
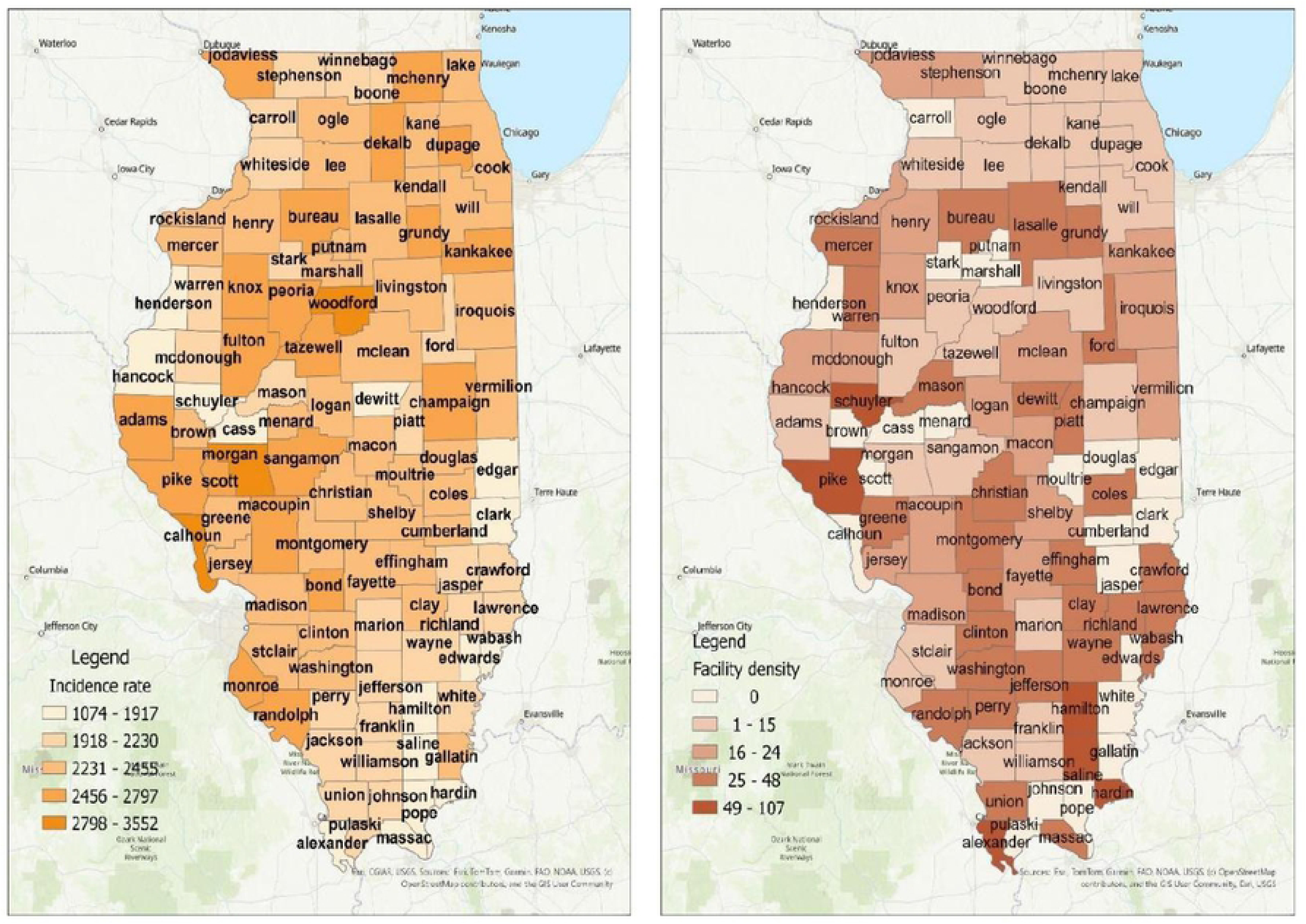
Choropleth maps of the incident rate and facility density across counties in Illinois.

**Table 1.** Descriptive statistics of incidence rate and facility density.

| Variables | Mean | SD | Median | Range | CV |
| --- | --- | --- | --- | --- | --- |
| Incidence rate | 2274.34 | 329.793 | 2297.33 | 2477.80 | 14.50% |
| Facility density | 19.00 | 17.94 | 15.39 | 107.41 | 94.39% |

Visual inspection of mapped surfaces suggested a loose correspondence between high incidence and high facility density in the state’s northern half but notable mismatches elsewhere, particularly in the rural south, where incidence remained moderate despite scant imaging resources (Fig 1).

A denominator sensitivity check confirmed that the observed gradients were not artifacts of population estimation. Replacing the five-year average female population with a single-year midpoint altered calculated incidence by less than five percent in 101 of 102 counties and never by more than ten percent, supporting the continued use of the averaged denominator for rate calculation (Table 2).

**Table 2.** Sensitivity analysis of average population (2017-2021) and midpoint population (2019)

| Category | Number of counties | % of counties |
| --- | --- | --- |
| Small (<5%) | 101 | 99.02% |
| Moderate (-10%) | 1 | 0.98% |
| Total | 102 | 100.00% |

Spatial statistics reinforced the impression of clustering. Global Moran’s I for breast cancer incidence equaled 0.27 (z = 4.99, p < 0.001), a moderate yet statistically robust signal of positive spatial autocorrelation. In contrast, Moran’s I for mammography density was –0.03 (p = 0.77), indicating no global spatial pattern for screening resources (Table 3). Local Indicator of Spatial Association analysis sharpened these results, revealing a continuous swath of high incidence running along the Illinois River valley through Peoria, Tazewell, Macoupin, and Pike counties. In contrast, a cluster of low incidence anchored the state’s far southern edge in Massac, Union, and White counties. On the other hand, low facility density appeared in scattered rural pockets without forming a coherent belt, implying that the disease burden is not confined to scanner-scarce locales (Fig 2; Table 4).

**Fig 2.**
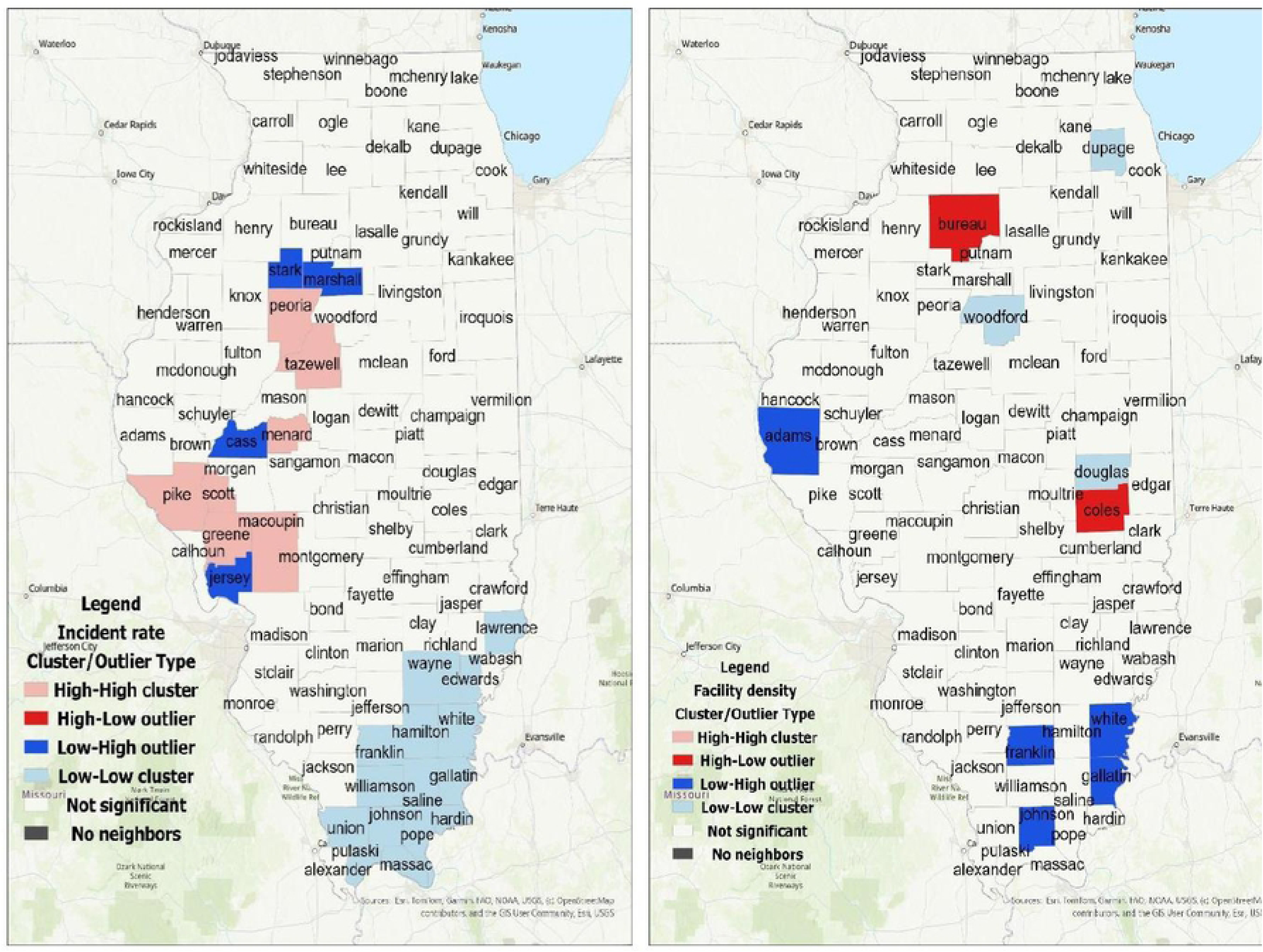
Choropleth maps of spatial clustering of the incident rate and facility density across counties in Illinois.

**Table 3.** Global Moran’s I Summary.

| **Feature** | **Incident Rate** | **Facility Density** |
| --- | --- | --- |
| Moran's I | 0.270 | -0.026 |
| Expected Index | -0.0099 | -0.0099 |
| Variance | 0.00314 | 0.00306 |
| Z-score | 4.998 | -0.299 |
| P-value | 0.000001 | 0.765 |
| Interpretation | Significant positive autocorrelation → clustered | No significant autocorrelation → random |
| Distance Band Used | 200,000 feet (≈ 38 miles) | 200,000 feet (≈ 38 miles) |

**Table 4.** Local Indicators of Spatial Association (LISA) summary.

|  | **Incident Rate** | **Facility Density** |
| --- | --- | --- |
| High-High cluster | Greene, Menard, Macoupin, Peoria, Pike, Scott, and Tazewell | None |
| High-Low Outlier | None | Bureau and Coles |
| Low-High outlier | Cass, Jersey, Marshall, and Stark | Adams, Franklin, Gallatin, Johnson, and White |
| Low-Low cluster | Edward, Franklin, Gallatin, Hamilton, Hardin, Johnson, Lawrence, Massac, Pope, Pulaski, Saline, Union, Wayne, White, and Williamson | Douglas and Woodford |
| Not significant | Others | Others |

Regression modeling began with an ordinary least-squares specification featuring facility density, bachelor’s attainment, poverty prevalence, and African American, Latino, and White proportions as covariates. This baseline model accounted for 17 percent of the between-county variance. A single percentage-point increase in bachelor’s degree attainment was associated with a 1,324-case rise per 100,000 women (p = 0.032), suggesting a detection effect tied to screening adherence in educated populations. Conversely, each additional mammography unit per 100,000 predicted a reduction of 4.91 cases (p = 0.007). Race proportions and poverty failed to reach conventional significance thresholds. However, diagnostic statistics indicated violations of OLS assumptions: the Jarque-Bera test rejected residual normality (p < 0.001), and residuals showed significant spatial clustering (Moran’s I = 0.19, p = 0.0005), pointing to model misspecification (Table 5).

**Table 5.** OLS Regression summary.

| Feature | Result |
| --- | --- |
| Model Type | Ordinary Least Squares |
| Dependent Variable | Breast Cancer Incidence Rate |
| Number of Observations | 102 |
| Adjusted R-squared | 0.173 |
| Akaike Information Criterion (AIC) | 1463.28 |
| Significant Predictors | Education Level (BACH_MIN), Facility Density |
| Education (Bachelor's or Higher) | $\beta = 1323.99$, $p = 0.032 \rightarrow$ Positive association |
| Facility Density | $\beta = -4.91$, $p = 0.0065 \rightarrow$ Negative association |
| Race Variables | African American, Latino, White → Not significant ($p > 0.3$) |
| Poverty Level | $\beta = -1453.25$, $p = 0.28 \rightarrow$ Not significant |
| Joint F-Statistic | $F(6,95) = 4.52$, $p = 0.0005 \rightarrow$ Model is statistically significant |
| Joint Wald Statistic | $\chi^2 = 39.29$, $p < 0.0001 \rightarrow$ Predictors jointly significant |
| Koenker (BP) Statistic | $\chi^2 = 4.60$, $p = 0.597 \rightarrow$ No heteroskedasticity detected |
| Jarque-Bera Test | $\chi^2 = 29.87$, $p < 0.0001 \rightarrow$ Residuals not normally distributed |

Incorporating a spatial-lag term markedly improved performance. The spatial coefficient (ρ = 0.49, p < 0.001) doubled the explained variance to 36.6 percent and lowered Akaike’s Information Criterion (AIC) by seventeen points relative to the OLS model. Education remained positive, and facility density was negatively associated with incidence (Table 6). Nevertheless, multicollinearity emerged as a serious problem among the three race variables: variance-inflation factors exceeded 25, pairwise Spearman correlations greater than 0.80, signaling instability (Fig 3).

**Fig 3.**
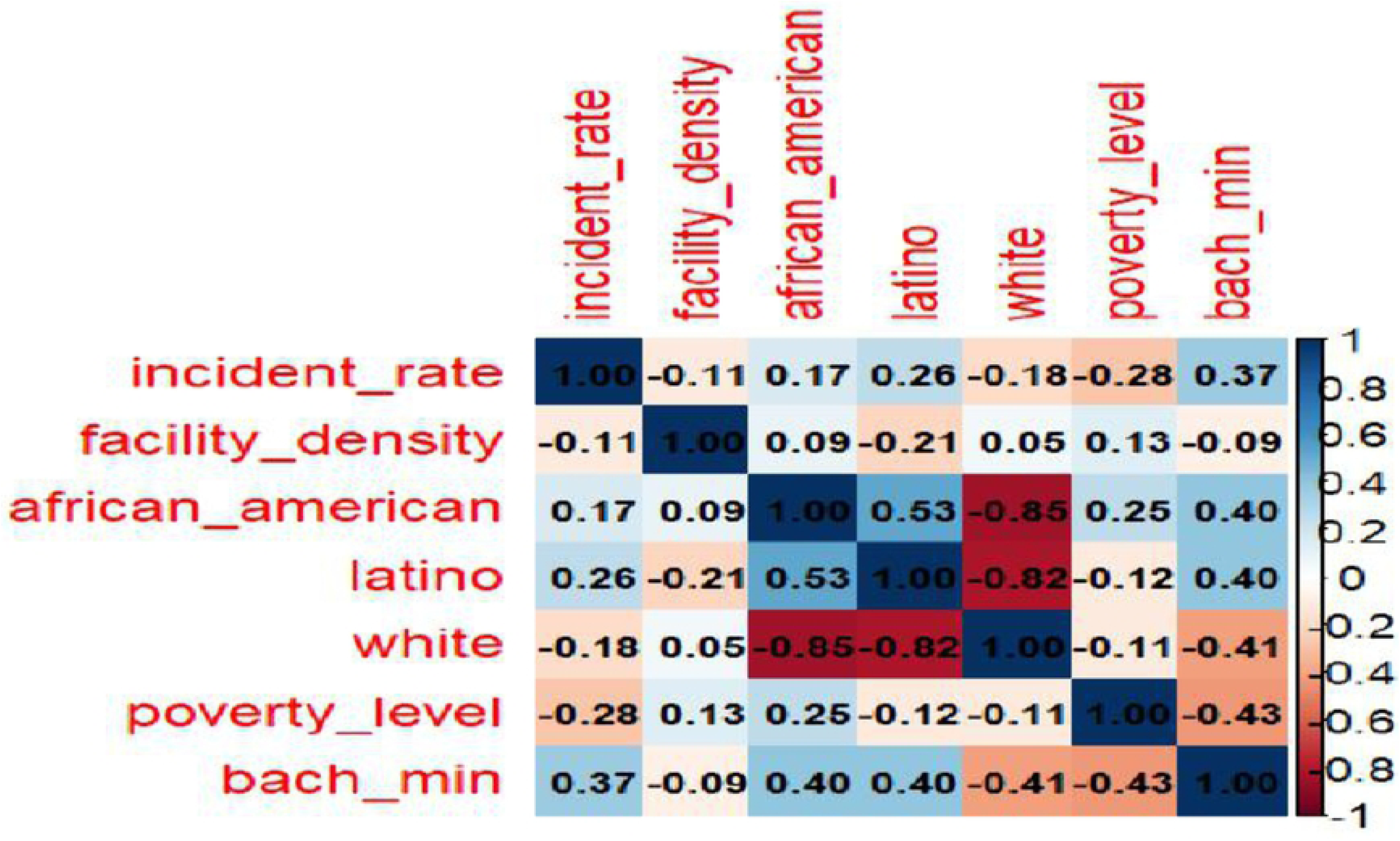
Spearman correlations linking breast cancer incidence, facility density, and sociodemographics across Illinois counties, 2017-2021.

**Table 6.** Spatial Lag model summary.

| Feature | Result |
| --- | --- |
| Model Type | Spatial Lag Model (Maximum Likelihood) |
| Dependent Variable | Breast Cancer Incidence Rate (T_Complete) |
| Number of Observations | 102 counties |
| Spatial Lag Coefficient ($\rho$) | 0.488 ($p < 0.001$) |
| R-squared | 0.366 (36.6% of variance explained) |
| Akaike Information Criterion (AIC) | 1446.38 |
| Significant Predictors | Education Level (T_Comple_5), Facility Density (T_Comple_6) |
| Education (Bachelor's or Higher) | $\beta = 1100.97$, $p = 0.047 \rightarrow$ Positive association |
| Facility Density | $\beta = -5.02$, $p = 0.001 \rightarrow$ Negative association |
| Race Variables | African American, Latino, White — Not significant ($p > 0.6$) |
| Poverty Level | $\beta = -435.28$, $p = 0.736 \rightarrow$ Not significant |
| Likelihood Ratio Test | $\chi^2 = 15.35$, $p = 0.00009 \rightarrow$ Model fits significantly better than OLS |
| Heteroskedasticity (Breusch-Pagan) | $\chi^2 = 10.37$, $p = 0.110 \rightarrow$ No evidence of heteroskedasticity (residuals OK) |

The three race proportions were reduced to a single principal component, capturing 96 percent of their combined variance to retain racial information without destabilizing estimates. A negative-binomial framework was adopted because incidence counts are over-dispersed relative to Poisson expectations (dispersion ratio ≈ 37). The PCA-adjusted negative-binomial model produced a residual deviance of 102.4 on 97 degrees of freedom and the lowest observed AIC (1,463). Under this specification, each additional mammography unit per 100,000 women corresponded to a 0.27 percent decline in incidence (β = –0.0027, p < 0.001). At the same time, bachelor’s degree attainment retained a positive association (β = 0.473, p = 0.022). The composite race index (p = 0.49) and poverty prevalence (p = 0.38) remained non-significant, suggesting that their apparent effects in simpler models stemmed from multicollinearity or confounding (Table 7).

**Table 7.** Comparison of Poisson and PCA-Adjusted Negative Binomial (NB) Regression Models for Breast Cancer Incidence Rate. The PCA-adjusted model was used to mitigate multicollinearity among race variables.

| Variable | Poisson Coef. (p-value) | PCA-NB Coef. (p-value) | Direction | Significant in NB? | Model Diagnostics |
| --- | --- | --- | --- | --- | --- |
| Intercept | 6.923 ($p < 0.001$) | 7.704 ($p < 0.001$) | + | Yes | Poisson AIC: ∞, NB AIC: 1462.6 |
| Facility Density | -0.0023 ($p < 0.001$) | -0.0027 ($p = 0.00043$) | - | Yes | Poisson Dev: 3832.2, NB Dev: 102.4 |
| Race Index (PCA) | — | -0.0111 ($p = 0.494$) | - | No | NB only |
| Poverty Level | -0.643 ($p < 0.001$) | -0.482 ($p = 0.379$) | - | No | |
| Bachelor's Degree | 0.574 ($p < 0.001$) | 0.473 ($p = 0.0215$) | + | Yes | |

Residual diagnostic supported the adequacy of the final model. A deviance residuals versus fitted plot showed a tight, pattern-free cloud centered on zero with no evidence of funneling, indicating homoscedasticity across the fitted range (Fig. 4). This diagnostic suggests that the negative-binomial specification successfully captured the data structure, leaving no systematic pattern in the residuals.

**Fig 4.**
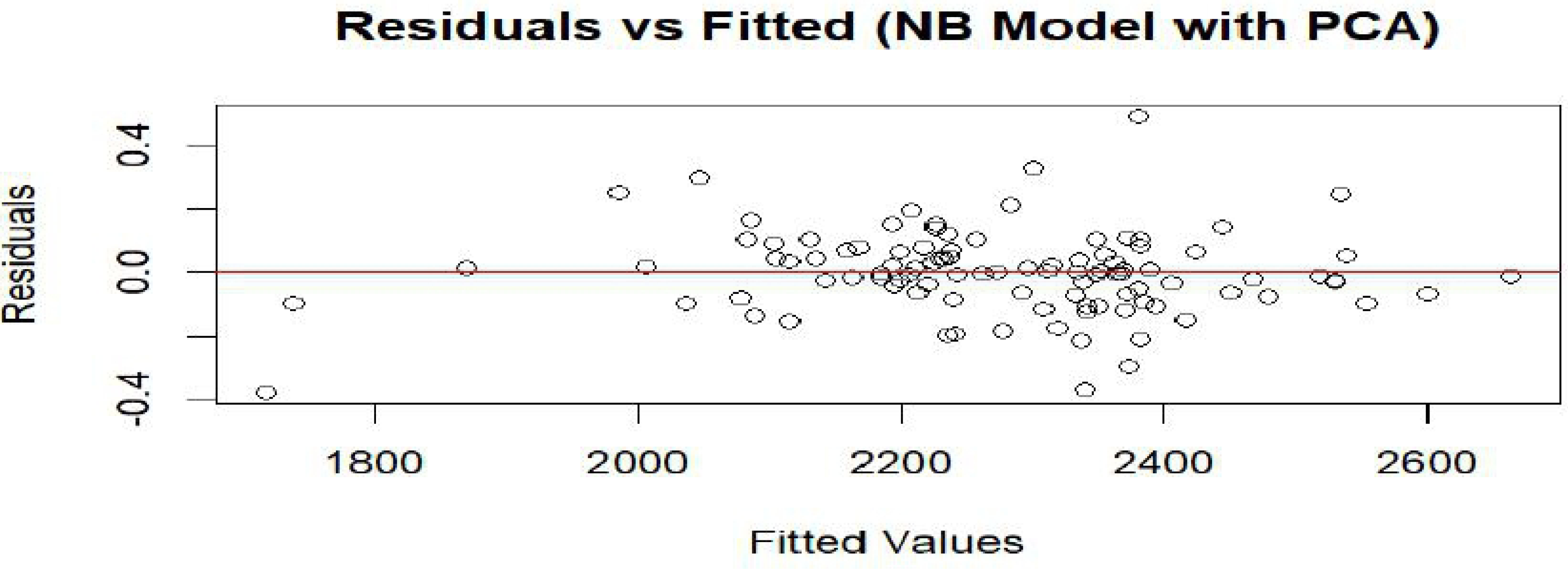
Scatter plot of deviance residuals versus fitted values from the PCA-adjusted Negative Binomial regression model.

Overall, spatial dependence and screening infrastructure explain a substantial share of the geographic variation in breast cancer incidence across Illinois. Facility density and educational attainment emerge as the dominant predictors after rigorous adjustment for spatial autocorrelation, over-dispersion, and multicollinearity. After structural and spatial elements are built into the model, neither the county’s racial mix nor its poverty rate shows an independent statistical link with breast-cancer incidence. However, many counties along the Illinois River still show unusually high case numbers, even though clinics and scanners are within easy reach. The fact that this hotspot persists suggests other forces may be at work, possibly something in the local environment or simply the pace at which women decide to get checked, and those possibilities need a closer look.

## Discussion

This study set out to map the uneven geographic distribution of breast cancer incidence and mammography access across Illinois and, in doing so, revealed complex structural, spatial, and social dynamics at play. While breast cancer remains a leading cause of morbidity and mortality among women in the United States, the underlying risk landscape is far from uniform. Our analysis, which leveraged spatial statistics and advanced regression modeling, highlights that geographic disparities are not simply random variations but reflect more profound systemic inequities that can influence when and whether women receive a diagnosis.

A key contribution of this research is the spatial framing of breast cancer burden in relation to healthcare infrastructure. The clustering of high-incidence counties, particularly in central Illinois, underscores that disease burden is not randomly distributed. The significant Moran’s I value for breast cancer incidence confirms the presence of spatial dependence, indicating that similarly burdened neighbors often surround counties with high rates. Such clustering suggests shared environmental, demographic, or healthcare access factors that may transcend administrative boundaries.

In contrast, mammography facility density exhibited no statistically significant spatial patterning, reflecting healthcare services’ disjointed and uneven planning. Prior studies have also reported the fragmented distribution of screening infrastructure, especially in rural and underserved regions, exacerbating delays in diagnosis and potentially contributing to disparities in outcomes [38,39]. This mismatch between where the burden lies and where resources are concentrated is a recurring theme in cancer control and prevention efforts [40].

Our regression models further elucidate these dynamics. Initially, OLS regression showed that educational attainment and facility density had statistically significant associations with breast cancer incidence. Notably, a higher percentage of women with at least a bachelor’s degree was linked to increased detection rates, likely reflecting better screening compliance or health-seeking behavior. Conversely, greater facility density was associated with slightly lower incidence rates, supporting the argument that accessibility can reduce diagnostic delay [41].

These results echo findings from similar U.S.-based research showing that social determinants such as education are critical facilitators of timely cancer screening [42].

However, the spatial lag model’s improved explanatory power—accounting for over 36% of the variation—suggests that traditional models without spatial terms may misestimate associations due to omitted spatial autocorrelation. This finding aligns with recent literature emphasizing the need for robust spatial-statistical methods in health geography to avoid biased inferences [43]. Our spatial lag coefficient (ρ = 0.49) was notably strong, implying that a county’s incidence rate is influenced by its own characteristics and neighboring counties. This spatial diffusion effect could stem from shared healthcare networks, regional referral patterns, or broader cultural norms regarding preventive health behavior.

The high multicollinearity among racial variables posed a methodological challenge, which we addressed using PCA. Multicollinearity inflates standard errors, making isolating the individual effects of variables like race and poverty difficult. By deriving a composite race index, we preserved interpretive power while avoiding the instability of overfit models. Though underutilized in epidemiologic studies, this technique is increasingly recommended for reducing redundancy in ecological regressions [44,45]. Even with PCA adjustment, race and poverty remained statistically non-significant in the final model, suggesting that their effects may be mediated through structural factors like education or spatially correlated contextual variables.

The transition from a Poisson model to a negative binomial framework was driven by the need to account for overdispersion, which can lead to underestimated standard errors and false inferences. This decision was supported by a dispersion ratio exceeding 30 and confirmed through formal statistical testing. The negative binomial model, particularly with PCA-adjusted covariates, yielded the best overall fit as indicated by lower deviance and AIC scores. This modeling approach has been validated in prior public health studies where count data exhibit more variability than expected under the Poisson assumption [46].

Notably, the final model’s findings emphasize that educational attainment and mammography facility density remain the most consistent predictors of county-level breast cancer incidence. This has direct implications for public health practice. Efforts to increase screening uptake should focus on expanding physical access to mammography centers and addressing informational and motivational barriers linked to education. Health literacy, often correlated with formal education, is key in shaping whether individuals engage in preventive care [47].

Moreover, despite adequate screening access, the persistence of high-incidence areas suggests that additional, perhaps less visible factors may be at play. These could include environmental exposures, genetic predispositions, or behavioral factors such as delayed help-seeking or mistrust in medical institutions. A growing body of research supports the notion that health outcomes are co-produced by social and environmental conditions rather than determined solely by access to services [48,49]. For Illinois specifically, examining agricultural chemical exposures or industrial pollutants in the high-incidence corridor could yield valuable insights.

The lack of significant findings for poverty in multivariate models warrants reflection. While poverty is a well-documented barrier to health services, its effects may be nonlinear or conditional based on other factors. Poverty’s impact might be attenuated in areas with high baseline access or substantial social capital. Conversely, in areas lacking infrastructure, poverty may compound risk in ways not fully captured by our models. Future studies incorporating interaction terms or mixed-effects designs could help unravel these nuanced dynamics [50].

The spatial analytic approach adopted here also has broader methodological implications. While much cancer epidemiology relies on aspatial techniques, this study demonstrates that spatial statistics can uncover patterns that would otherwise remain hidden. Integrating LISA maps with regression diagnostics allows for a more nuanced understanding of public health landscapes and can inform targeted interventions. Policymakers and program planners can use such findings to prioritize counties where structural barriers coincide with elevated disease burden.

This study does have limitations. As an ecological analysis, it cannot account for individual-level behaviors or exposures, raising the possibility of ecological fallacy. Moreover, our measure of facility density does not capture appointment availability, travel times, or perceived quality factors known to influence screening behaviors. While we geocoded facility locations, our analysis did not incorporate transportation infrastructure or insurance networks, which could further explain disparities. Nonetheless, the consistency of our findings across multiple analytic strategies lends confidence to our core conclusions.

This research advances our understanding of Illinois’s spatial determinants of breast cancer detection. By combining ecological modeling, spatial analysis, and dimensionality reduction, we comprehensively portray how geography, infrastructure, and sociodemographic context intersect to shape health outcomes. These insights are particularly salient for states like Illinois, encompassing diverse urban, suburban, and rural communities. Moving forward, health equity efforts should consider who has access and where and how that access is distributed.

## Conclusion

Despite modest progress in cancer control, significant geographic disparities in breast cancer outcomes remain embedded within Illinois’s healthcare landscape. This study shows that incidence rates and screening infrastructure are unevenly distributed, with some counties experiencing disproportionate burdens despite appearing adequately resourced. The evidence points to deeper, systemic challenges, where the mere presence of facilities does not guarantee equitable access or use. Education and spatial clustering emerged as strong drivers of variation, while traditional sociodemographic predictors like race and poverty lost statistical power after controlling for spatial autocorrelation and multicollinearity.

Closing the gap between underserved and over-resourced areas demands multifaceted policy solutions. First, expanding mobile mammography units has successfully reduced screening disparities, particularly in rural areas. For example, mobile clinics in South Carolina and West Virginia increased mammography uptake among uninsured and underinsured women, bringing services directly to hard-to-reach communities [51]. These mobile programs are logistically feasible in large, geographically diverse states like Illinois and could be strategically deployed in counties identified as low-access clusters through spatial mapping.

Second, transportation remains an often-overlooked barrier. Non-emergency medical transportation (NEMT) services funded through Medicaid have shown promise. However, recent studies suggest that rideshare partnerships, such as those piloted by Philadelphia and Los Angeles clinics, offer scalable, cost-effective alternatives to reduce missed appointments [52]. Embedding such services within existing community health initiatives could lower logistical hurdles for women in car-dependent or transit-poor counties.

Third, patient navigation programs that pair women with trained navigators from their communities have successfully enhanced follow-up after abnormal screening and increased adherence to recommended diagnostic pathways. Peer navigator interventions, particularly those that are culturally tailored, have effectively addressed distrust, language barriers, and informational gaps in historically marginalized populations [53]. Scaling up navigator programs in high-incidence but high-access areas, where other structural issues persist, could make a substantial difference.

State and local health departments should consider allocating targeted funding based on geospatial burden profiles. Counties identified as "high–high" clusters with elevated incidence and proximity to similar counties warrant prioritized intervention funding and enhanced surveillance to explore environmental or healthcare behavior factors. Such targeting would increase efficiency and address equity by accounting for spatial spillover effects, as demonstrated in our spatial lag models.

Finally, future research should extend beyond facility counts and ecological correlates to include individual-level screening histories, insurance coverage, travel time estimates, and perceived barriers. Integrating electronic health record data with geographic information systems (GIS) may uncover actionable insights to refine public health strategies.

Illinois’s geographic diversity, spanning urban centers to rural farmland, requires flexible, community-driven care delivery models. By combining spatial analysis with targeted intervention design, public health stakeholders can begin to bridge the persistent divide between resource availability and population outcomes.

## Data Availability

All supporting data are available from the https://doi.org/10.5281/zenodo.15420318. All relevant data are also available within the manuscript and its supporting information files.

## Supporting information

**S1 Data. Data dictionary and analysis datasets (XLSX)**

**S1 Text. Comprehensive analytical workflow (PDF)**

**S2 Text. R script and statistical results (PDF)**

**S3 Text. GeoDa spatial analysis output (PDF)**

**S4 Text. Illinois mammography facilities data (PDF)**

**S1 File. ArcGIS project files (ZIP)**

**S2 File. US Census demographic data (ZIP)**

**S3 File. Illinois County Shapefiles (ZIP)**

